# Gaussian Statistics and Data-Assimilated Model of Mortality due to COVID-19: China, USA, Italy, Spain, UK, Iran, and the World Total

**DOI:** 10.1101/2020.04.06.20055640

**Authors:** T.-W. Lee, J.E. Park, David Hung

## Abstract

Covid-19 is characterized by rapid transmission and severe symptoms, leading to deaths in some cases (ranging from 1.5 to 12% of the affected, depending on the country). We identify the Gaussian nature of mortality due to covid-19, as shown in China where it appears to have run its course (during the first sweep of the pandemic at least) and other coutnries, and also in Imperial College modeling. Gaussian distribution involves three parameters, the height, peak location and the width, and the streaming data can be used to infer function value, slope and inflection location as a minimum set of constraints to estimate the subsequent trajectories. Thus, we apply the Gaussian function template as the basis for a data-assimilated model of covid-19 trajectories, first to USA, United Kingdom (UK), Iran and the world total in this study. As more data become available, the Gaussian trajectories are updated, for other nations and also for state-by-state projections in USA.

## INTRODUCTION

Covid-19 is characterized by rapid transmission and severe symptoms, leading to deaths in some cases (ranging from 1.5 to 12% of the affected, depending on the country [1]). Due to the pandemic nature and severity, it is essential to do the maximum possible in all areas of response, control and mitigation. This primarily needs to occur in the medical field, in terms of patient care, treatment and vaccine developments. However, in order to organize and prepare medical responses, modeling and prediction of Covid-19 transmission and its outcome are also important. The Imperial College study has been timely and instrumental in this regard [2], and we also use their results as one of the supporting data for current work. In this study, we identify the Gaussian nature of mortality due to Covid-19, as shown in China where it appears to have run its course (during the first sweep of the pandemic at least) and other coutnries, and also in Imperial College modeling [2]. We apply the Gaussian statistical analyses to USA, United Kingdom (UK), Iran and the world total in this first study. Gaussian distribution involves three parameters, the height, peak location and the width. For a scattered data set, there is an infinite number of permutations of these parameters. However, for Gaussian distribution even with limited data sets, there are several inflection points, which can be used as milestone for guiding the trajectory of the Gaussian function. For example, first, second, and third gradients of the Gaussian probability distribution function are plotted in Fig. 1. The first gradient is of course the slope, the second gradient curvature, and so on. We can see that the first inflection point where the Gaussian function starts to rise steeply occurs on Day 33 in this example. Therefore, even with data sets with scatter, using the least-fits for the function value, slope and the inflection point furnish us with minimum necessary conditions to estimate the particular Gaussian function. As further data become available and further inflection points (particularly the peak location) are identified, then rapid convergence toward reasonably accurate Gaussian estimate becomes feasible. This of course is an idealized situation, and real data include scatter, which makes it difficult to evaluate even the gradient with desired accuracy. Nonetheless, we can still estimate the upper and lower bounds of the trajectories, by using the Gaussian probability density function as a template. As more data stream in, the accuracy of the Gaussian “model” is continuously improved. If the second inflection point (the peak in the Gaussian distribution) can be identified from the data, as in the case of China, Italy, and Spain (based on the data thus far, as of April 6, 2020), then substantial improvements in the overall Gaussian model can be attained. This however becomes more or less retrospective analysis by that point. This work is aimed at achieving upper- and lower-bound projections of current and subsequent trajectories of Covid-19, or similar pandemic spread in the future, based on Gaussian nature of mortality statistics.

**Fig. 1.**
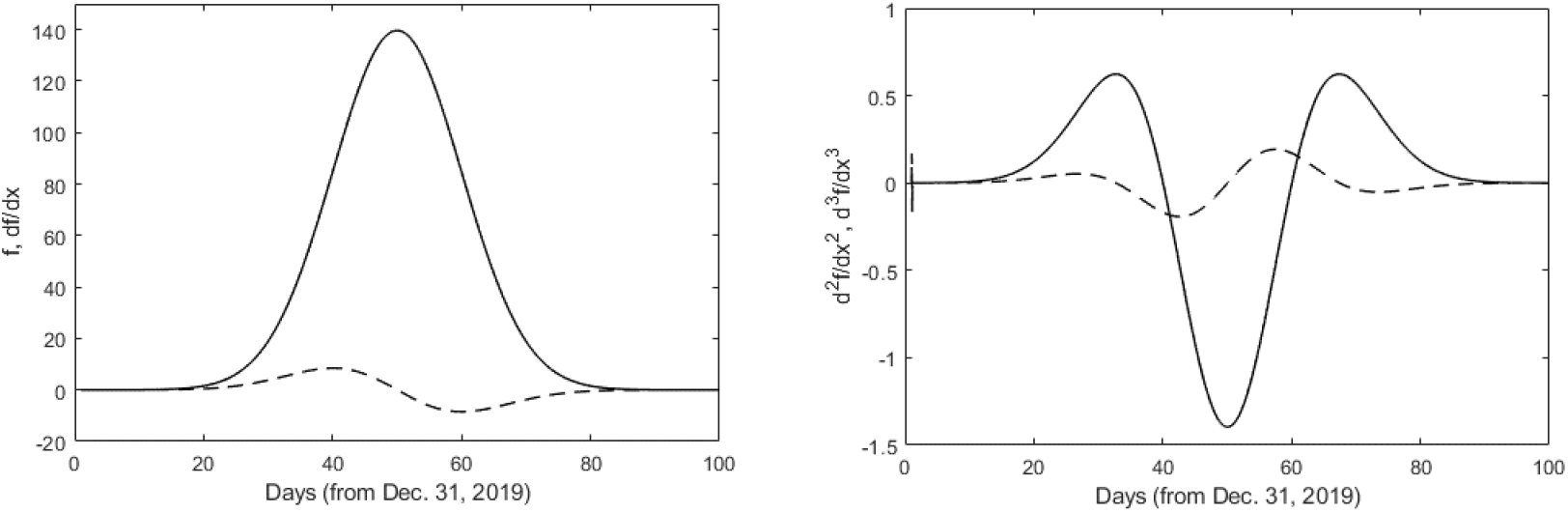
Characteristics of Gaussian distribution function, including its gradients.

### STATISTICAL METHOD

The mortality statistics from China, Italy and Spain are shown in Figs. 2(a), (b) and (c). The data are from Ref.1, and current Gaussian fits are plotted as lines. These three nations, particularly China, exhibit statistics that are further developed in time (as of April 6, 2020), thus showing the overall trend. The important inflection points are present in these statistics. Initially gradual increase is marked by a sudden expansion, resembling an exponential growth. Then, leveling occurs at the peak, followed by a descent. We can see that in spite of some scatter, the trends follow Gaussian or normal distribution in China, Italy and Spain. This appears to be a common attribute of mortality statistics, where the modeling study by the Imperial College group [2] also shows the Gaussian distribution for both the mortality rate (Fig. 3) and number of required critical beds, where for the latter the width and height parameters differ when intervention methods are applied but the statistical form is retained [2]. As noted in the introduction, we exploit this trait by using the Gaussian template with three statistical function parameters, A, m and, as expressed in Eq. 1.

**Fig. 2.**
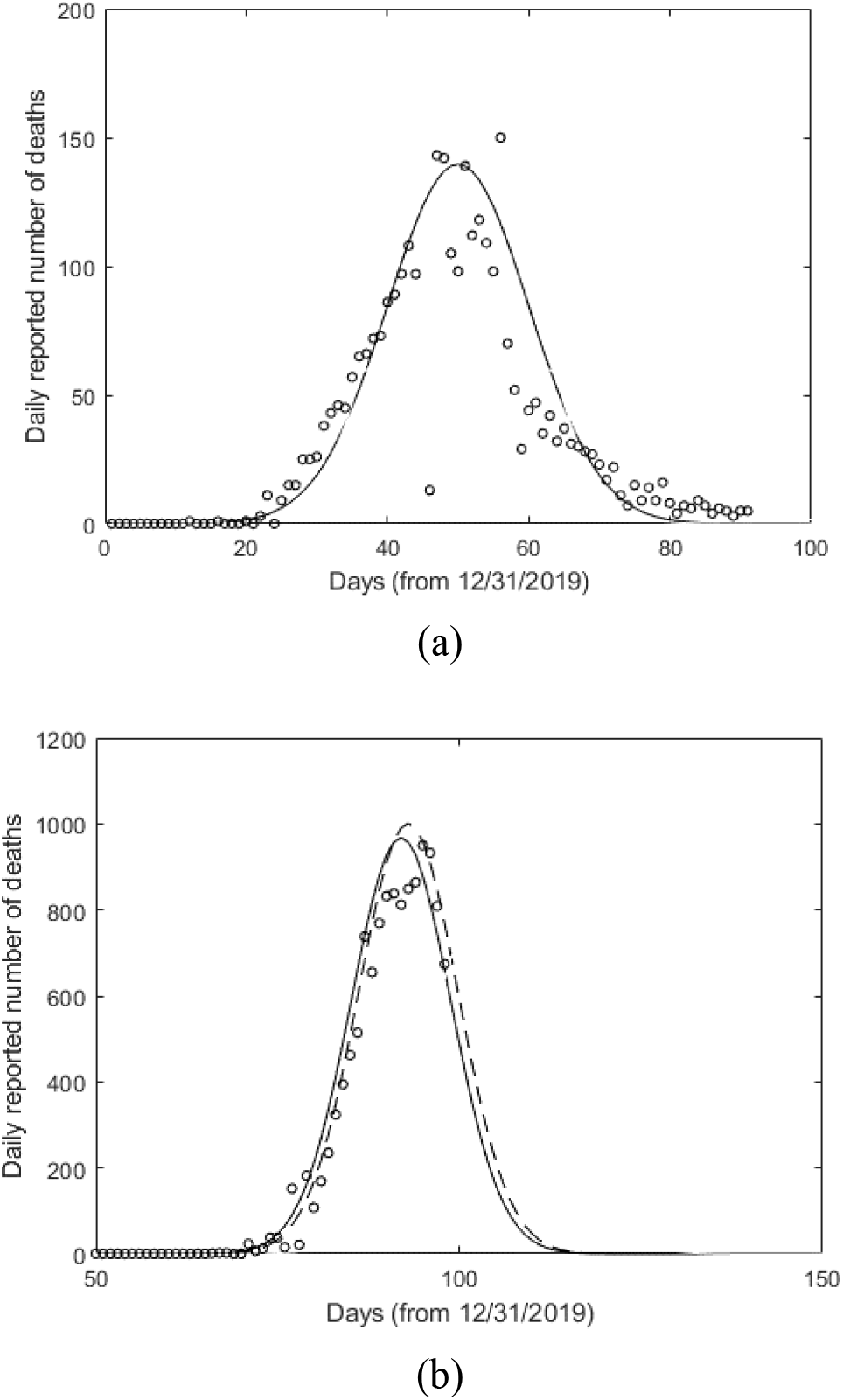
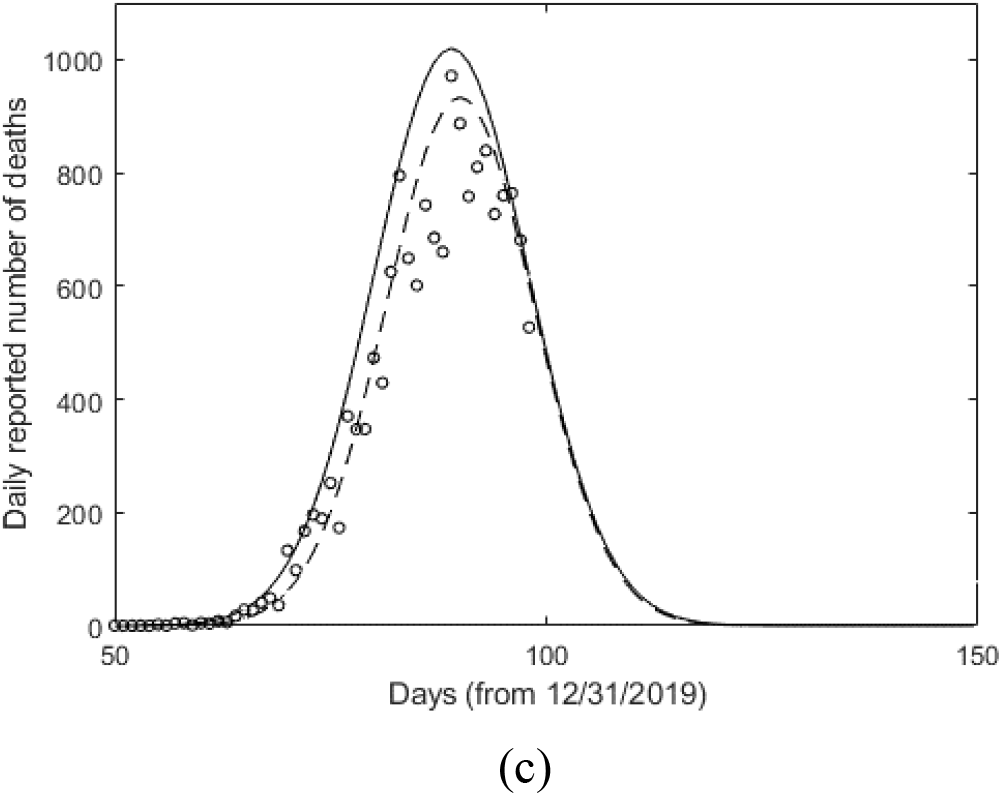
Statistical data (circles) and Gaussian model (line) for China (a), Italy (b) and Spain (c).

**Fig. 3.**
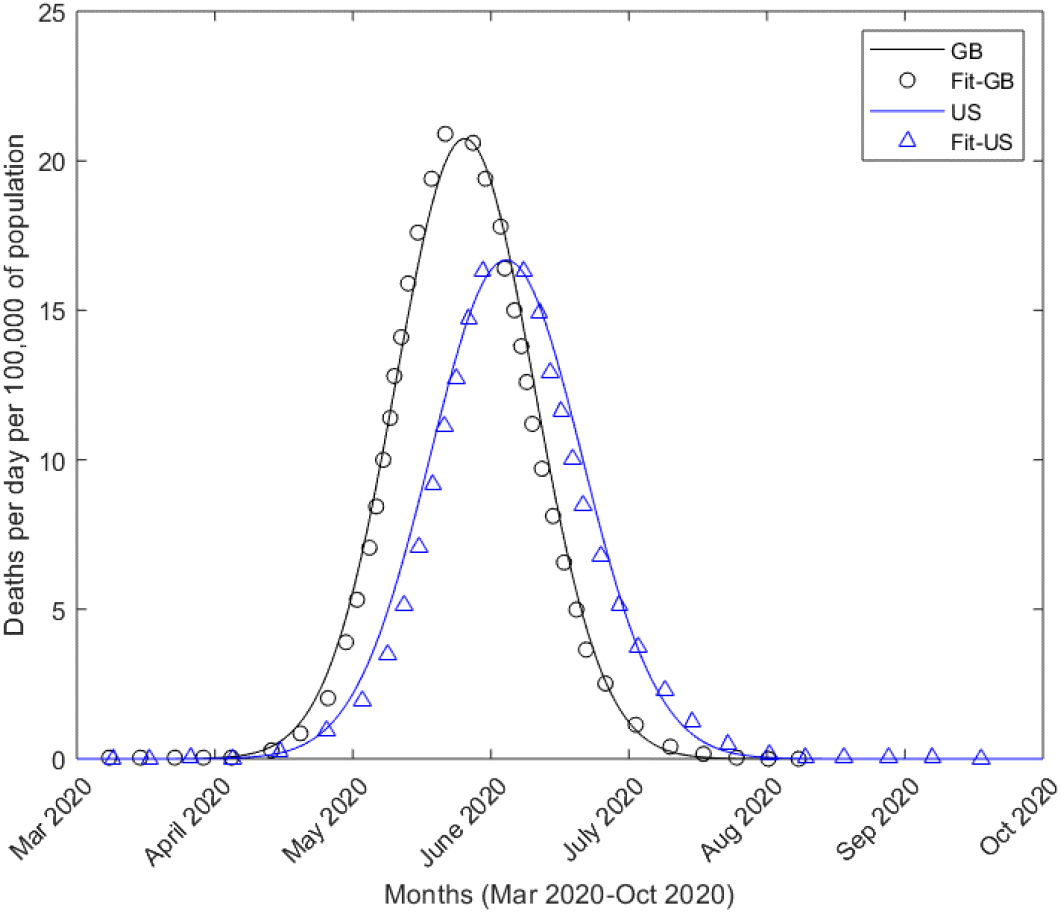
Comparison of Imperial College model results (symbols) with Gaussian fits (lines) for USA and UK (GB).

The function parameters in Eq. 1 can be progressively optimized using the least-square fit with data that reproduces the function value, slope and the location of the first inflection point (d^3^f/dx^3^ = 0). Due to the nonlinear nature of f(x), the three conditions may produce multiple solutions (possibilities) for the final f(x). However, this is expected to doublets or at most triplets with some non-physical solutions, and thus this approach is far more preferable than having to deal with an infinite number of permutations of in function parameters. In any event, since the data involves scatter and a short time-duration, a unique solution for f(x) is very difficult to achieve, if not impossible, in any useful and timely fashion. Depending on the level and timing of intervention, the function parameters also may shift, although thus far due to possible ergodic nature of the data the statistics exhibit nearly fixed Gaussian shape within the error bounds. For these reasons, we focus on “inner” and “outer” trajectory limits, which will lead to upper and lower estimates of the total mortality. These trajectories and estimates are continuously optimized using streaming data (2 times/week), as this work is being reviewed.

## RESULTS AND DISCUSSION

Figs. 4-8 are the results of above statistical estimates of mortality trajectories for Spain, USA, UK, Iran and the world total. They all show the beginning phase of Gaussian distribution function, and above method is applied to obtain inner and outer trajectories, in order to account for scatter in the data. As noted above, data from the initial phase including the first inflection point including the number and slope at that point are the minimum requirements for Gaussian trajectory estimates. Two Gaussian fits are plotted as lines, “inner” as solid and “outer” as broken lines. Iran is a different case than others, where there appears to be a beginning of a second peak, in which case it may become necessary to use two overlapping Gaussian functions. Also, UK is a difficult case to assess due to spikes during the initial phase. Based on these estimates thus far, the peak and total mortality can be estimated. As this work goes under review, and possibly publication, the data will be updated along with updated estimates for the peak and total mortality data through off- and online sites. In addition, other nations showing substantial statistics of covid-19 will be analyzed, followed by state-by-state analyses of USA.

**Fig. 4.**
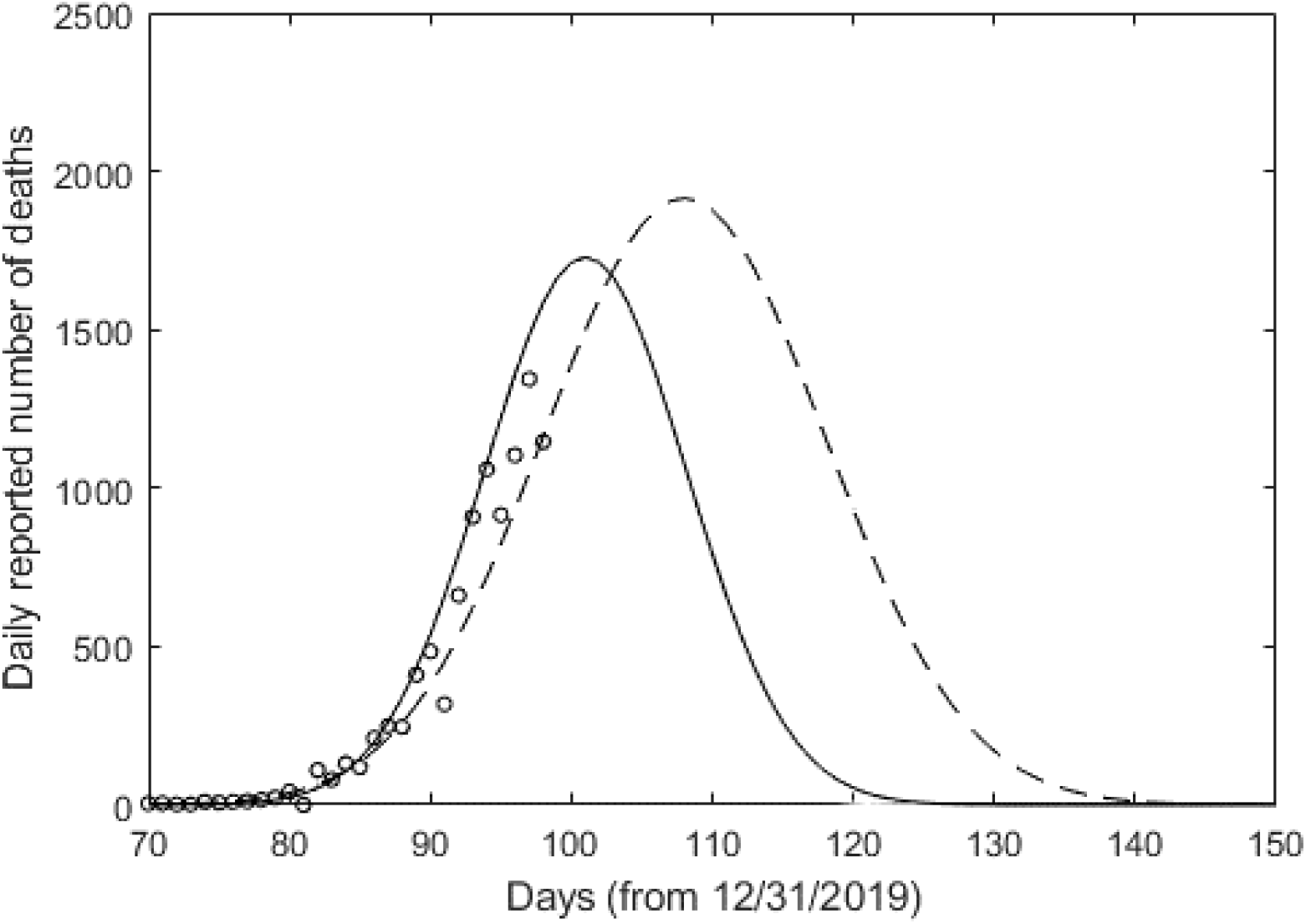
Statistical data (circles) and Gaussian model (line) for USA.

**Fig. 5.**
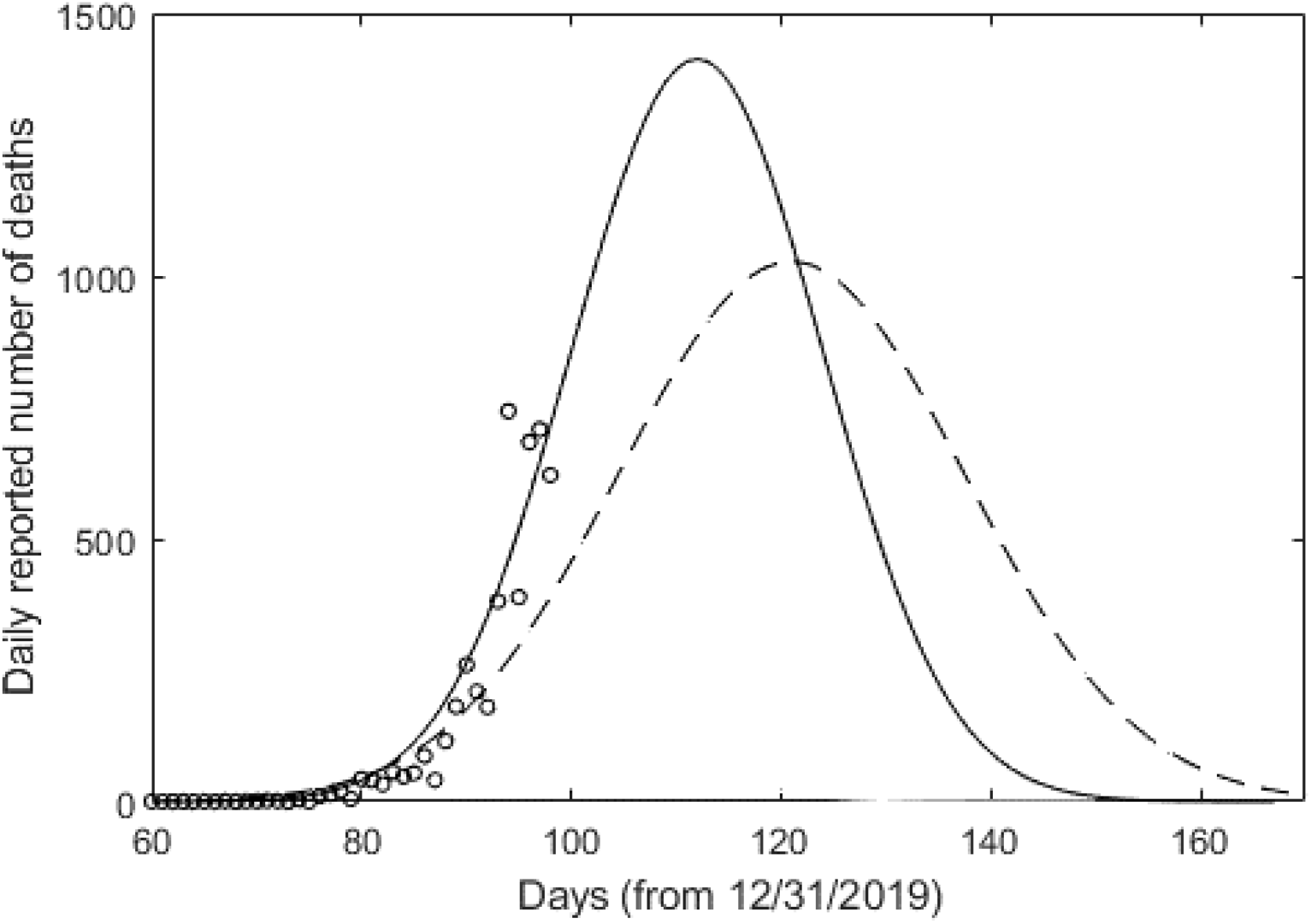
Statistical data (circles) and Gaussian model (line) for United Kingdom.

**Fig. 6.**
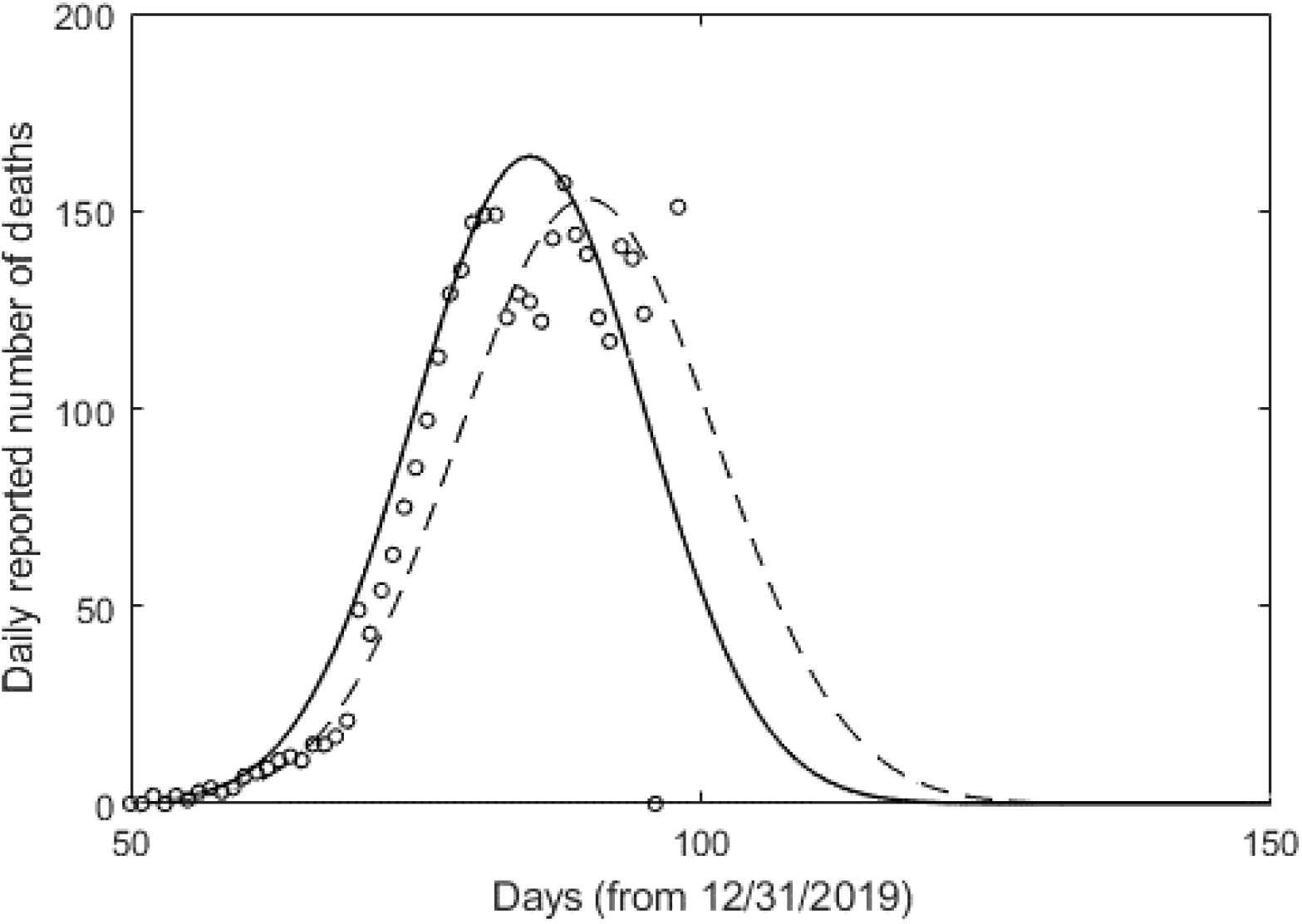
Statistical data (circles) and Gaussian model (line) for Iran.

**Fig. 7.**
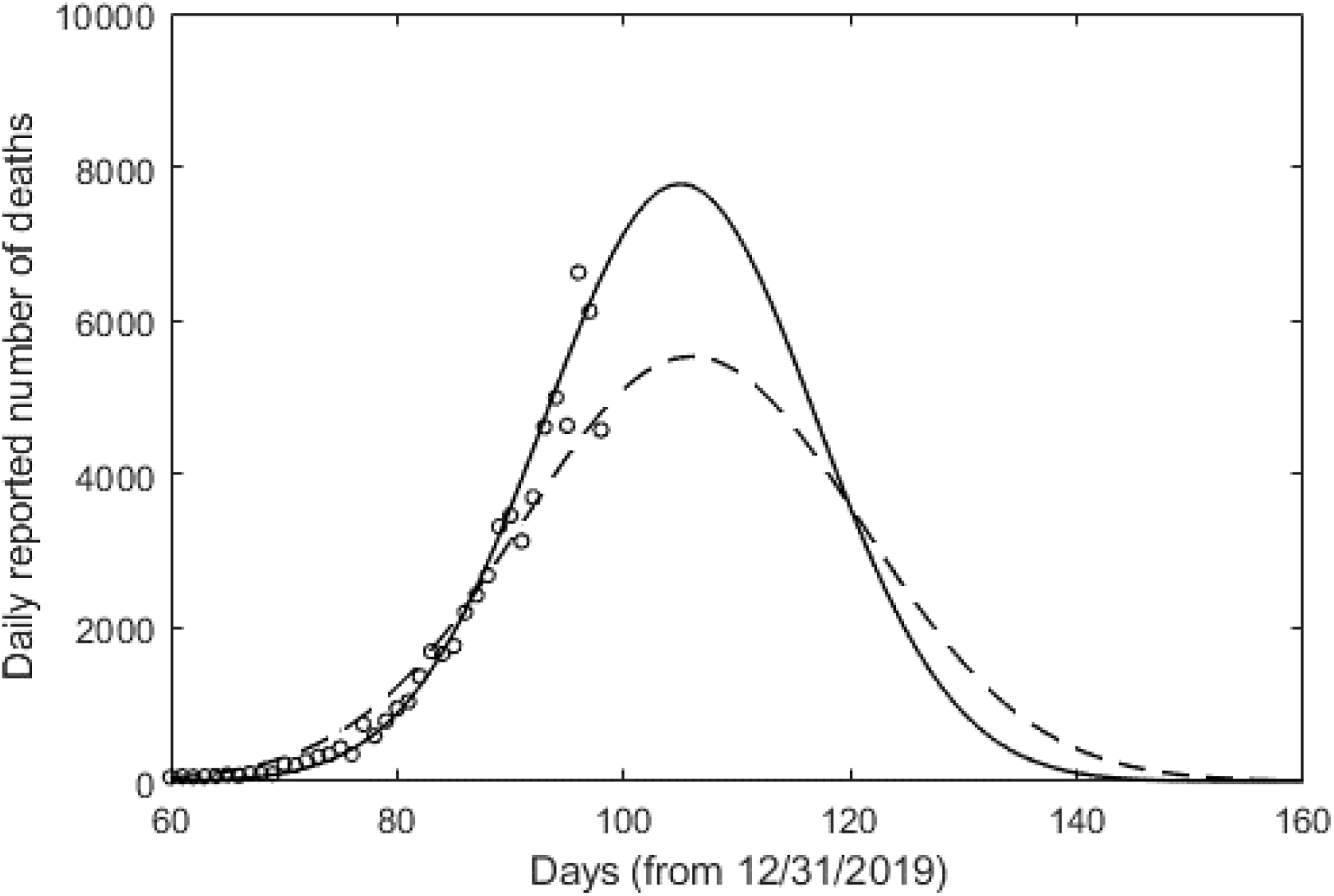
Statistical data (circles) and Gaussian model (line) for the World total.

## Data Availability

The data have been obtained from a public source (Ref 1).
Our own analysis data are available upon request, and will be posted online after a website is set up.

